# An Interpretable Machine Learning Framework for Accurate Severe vs Non-severe COVID-19 Clinical Type Classification

**DOI:** 10.1101/2020.05.18.20105841

**Authors:** Yuanfang Chen, Liu Ouyang, Forrest Sheng Bao, Qian Li, Lei Han, Baoli Zhu, Ming Xu, Jie Liu, Yaorong Ge, Shi Chen

**Affiliations:** Public Health Research Institute of Jiangsu Province, Nanjing, 210009, China; Institute of HIV/AIDS/STI Prevention and Control, Jiangsu Provincial Center for Disease Control and Prevention, Nanjing, 210009, China; Department of Orthopedics, Union Hospital, Tongji Medical College, Huazhong University of Science and Technology, Wuhan, 430022, China; Department of Computer Science, Iowa State University, Ames, IA, 50011, USA; Department of Pediatrics, Affiliated Kunshan Hospital of Jiangsu University, Kunshan, 215300, China; Department of Occupational Disease Prevention, Jiangsu Provincial Center for Disease Control and Prevention, Nanjing, 210009, China; School of Public health, Nanjing Medical University, Nanjing, 211166, China; Department of Public Health Sciences, College of Health and Human Services, University of North Carolina Charlotte, Charlotte, NC 28223, USA; Department of Radiology, Union Hospital, Tongji Medical College, Huazhong University of Science and Technology, Wuhan, 430022, China; Department of Software and Information Systems, College of Computing and Informatics, University of North Carolina Charlotte, Charlotte, NC 28223, USA; School of Data Science, University of North Carolina Charlotte, Charlotte, NC 28223, USA

## Abstract

Effectively and efficiently diagnosing COVID-19 patients with accurate clinical type is essential to achieve optimal outcomes for the patients as well as reducing the risk of overloading the healthcare system. Currently, severe and non-severe COVID-19 types are differentiated by only a few clinical features, which do not comprehensively characterize complicated pathological, physiological, and immunological responses to SARS-CoV-2 invasion in different types. In this study, we recruited 214 confirmed COVID-19 patients in non-severe and 148 in severe type, from Wuhan, China. The patients’ comorbidity and symptoms (26 features), and blood biochemistry (26 features) upon admission were acquired as two input modalities. Exploratory analyses demonstrated that these features differed substantially between two clinical types. Machine learning random forest (RF) models using features in each modality were developed and validated to classify COVID-19 clinical types. Using comorbidity/symptom and biochemistry as input independently, RF models achieved >90% and >95% predictive accuracy, respectively. Input features’ importance based on Gini impurity were further evaluated and top five features from each modality were identified (age, hypertension, cardiovascular disease, gender, diabetes; D-Dimer, hsTNI, neutrophil, IL-6, and LDH). Combining top 10 multimodal features, RF model achieved >99% predictive accuracy. These findings shed light on how the human body reacts to SARS-CoV-2 invasion as a unity and provide insights on effectively evaluating COVID-19 patient’s severity and developing treatment plans accordingly. We suggest that symptoms and comorbidities can be used as an initial screening tool for triaging, while biochemistry and features combined are applied when accuracy is the priority.

**One Sentence Summary:** We trained and validated machine learning random forest (RF) models to predict COVID-19 severity based on 26 comorbidity/symptom features and 26 biochemistry features from a cohort of 214 non-severe and 148 severe type COVID-19 patients, identified top features from both feature modalities to differentiate clinical types, and achieved predictive accuracy of >90%, >95%, and >99% when comorbidity/symptom, biochemistry, and combined top features were used as input, respectively.

## Introduction

COVID-19 is a pandemic caused by the novel SARS-CoV-2 virus. As of May 17 2020, it has spread through at least 220 countries and regions, causing more than 4 million cases with 300 thousand casualties (1). It is considered as the single most severe outbreak in the entire world during the 21st century, dwarfing other coronavirus-caused 2003 SARS and 2012 MERS epidemics. COVID-19 is especially challenging to the health professionals and general population. Unlike the precedent SARS and MERS epidemics, COVID-19 patients can be either asymptomatic or symptomatic, both of which are demonstrated to be transmissible of the virus with varying degrees (2-5). In addition, the distinct clinical types, non-severe and severe, require different treatment and care plans (6). Current studies are able to differentiate COVID-19 patients from non-patients, but further detecting non-severe or severe types of COVID-19 is not comprehensively explored. Non-severe type patients can be accommodated in the mobile cabin hospital which requires relatively less intensive clinical monitoring and intervention, including treating pre-existing comorbidities, preventing healthcare associated infections and other comorbidities (8). In contrast, severe type patients need close monitoring, usually in ICU with more clinicians (6). Therefore, effectively and efficiently classifying COVID-19 clinical types is essential for triage, resource optimization, and care planning for front-line clinicians, healthcare systems, as well as for the patients (6,7).

Currently, non-severe and severe type are classified based on only a few clinical features (shortness of breath, O_2_ saturation, and PaO_2_), which do not comprehensively characterize the complicated pathological, physiological, and immunological profile between non-severe and severe types in COVID-19 patients (9-11). In addition, some severe patients may not present shortness of breath initially. But without proper medical intervention, their clinical course will worsen abruptly, often resulting in respiratory failure with high mortality (6). It is therefore critical to provide accurate and efficient diagnosis of COVID-19 patients with correct clinical type information. We suggest that clinical features, including patient’s comorbidities (e.g., hypertension and diabetes), clinical symptoms (e.g., fever and chest pain), and blood biochemistry, are able to provide a more comprehensive characterization of COVID-19 and differentiate its clinical types (12,13). The human body is a unified and integrated entity. When pathogens such as SARS-CoV-2 invade, its effects can be shown not only from CT scans in the thoracic region, but also from other aspects such as clinical symptoms and biochemistry. ACE-2 receptors, which facilitate SARS-CoV-2 infiltration, are distributed across multiple organs and systems in human body (35). More recent discoveries have found that in addition to respiratory system, SARS-CoV-2 can also invade digestive, reproductive, and even neural systems as well (14-17). In other words, comorbidities, clinical symptoms, and blood biochemistry information of COVID-19 patients could all be consequences and/or risk factors of SARS-CoV-2 infection.

In clinical practice against COVID-19, clinicians not from respiratory or intensive care units may rely only on the referenced symptoms and signs (9) while neglecting diverse and important clinical features of COVID-19 patients, and may miss the critical signs of clinical course, leading to undesirable clinical consequences.

The potential power of symptoms, blood biochemistry, as well as their combinations to determine COVID-19 clinical type is currently not well understood nor evaluated (18-20). In order to utilize such diverse multimodality clinical information to make accurate and interpretable classifications, we propose a data mining and machine learning (ML) framework alternative to commonly used hypothesis-driven parametric models such as logistic regression. The results can provide reliable diagnostic decision support for clinicians even without comprehensive experience on the emerging COVID-19. We aim to explore and contrast the distributions of comorbidities and symptoms, as well as blood biochemistry between non-severe and severe COVID-19 types. We will identify key features that differed substantially between the two clinical types and provide clear evidence-based interpretations for clinicians and other health professionals. Next, we will investigate whether single modality or specific combination of features across modalities are able to provide accurate classification models based on ML techniques. This study delivers an accurate diagnostic decision support tool to differentiate non-severe from severe type patients based on commonly available clinical data with clear clinical interpretations. Insights gained from this study, as well as developed end-to-end multimodal data analysis and ML framework, will enable us to better understand the comprehensive pathology of COVID-19, further distinguish COVID-19 from other infectious respiratory diseases, and apply in other diseases with multimodal clinical data in the future.

## Results

### Data Mining of COVID-19 Clinical Features between Non-severe and Severe Types

Prevalence of symptom features in non-severe and severe COVID-19 types were calculated and compared (Fig. 1). Patients in the two clinical types showed distinct prevalence of many features. Severe COVID-19 patients were statistically much more likely to be elderly (at least age of 50, symbol OLD, OR=13.77, 95% CI= 7.33-25.86, p<0.001) and male (SEX, OR=1.89, 95% CI= 1.24-2.90, p<0.01), to have renal diseases (KID, OR=8.51, 95% CI= 1.86-38.99, p<0.001), cardiovascular diseases (CAR, OR=5.61, 95% CI=2.81-11.20, p<0.001), hypertension (HYP, OR=5.37, 95% CI=3.36-8.56, p<0.001), diabetes (DIA, OR=4.61, 95% CI=2.53-8.38, p<0.001), loss of appetite and taste (NAP, OR=3.20, 95% CI=1.70-6.01, p<0.001), feeling chilly (CHL, OR=2.21, 95% CI=1.16-4.22, p<0.05), and chest congestion (SHB, OR=1.88, 95% CI=1.22-2.89, p<0.01) than their non-severe counterparts. The only exception was sore throat, where severe patients had significantly much less likelihood to develop (SOR, OR=0.30, 95% CI=0.14-0.61, p<0.001). Since some of these clinical conditions were self-reported symptoms, observation biases were to be expected. These discoveries were further demonstrated in the forest plot of odds ratio (OR) and CI in Fig. 2, showing the differences between the two clinical types. Therefore, these relatively easily measured and acquired clinical features could be utilized to clinically evaluate COVID-19 patients’ severity. Our findings also echoed the U.S. CDC’s recently updated list of symptoms of COVID-19 (21) and more recent reports on characterizations of COVID-19 patients in the U.S. (36). Our findings showed that elderly male COVID-19 patients with cardiovascular, respiratory, renal diseases and diabetes were at much higher risk of developing serious complications of COVID-19 such as acute respiratory distress syndrome (ARDS) and even death (19,20). In addition, we discovered that Chinese patients with renal diseases were significantly more likely to develop severe COVID-19, which was not widely reported before. Clinical evidence showed that ACE-2 expression was associated with kidney diseases, thus making kidney disease a potential complication of SARS-CoV-2 invasion (22,23). This finding would inform clinicians to monitor kidney dysfunction, e.g., acute kidney injury, as a clinical sign and/or consequence of severe COVID-19 complication as well.

**Fig. 1.**
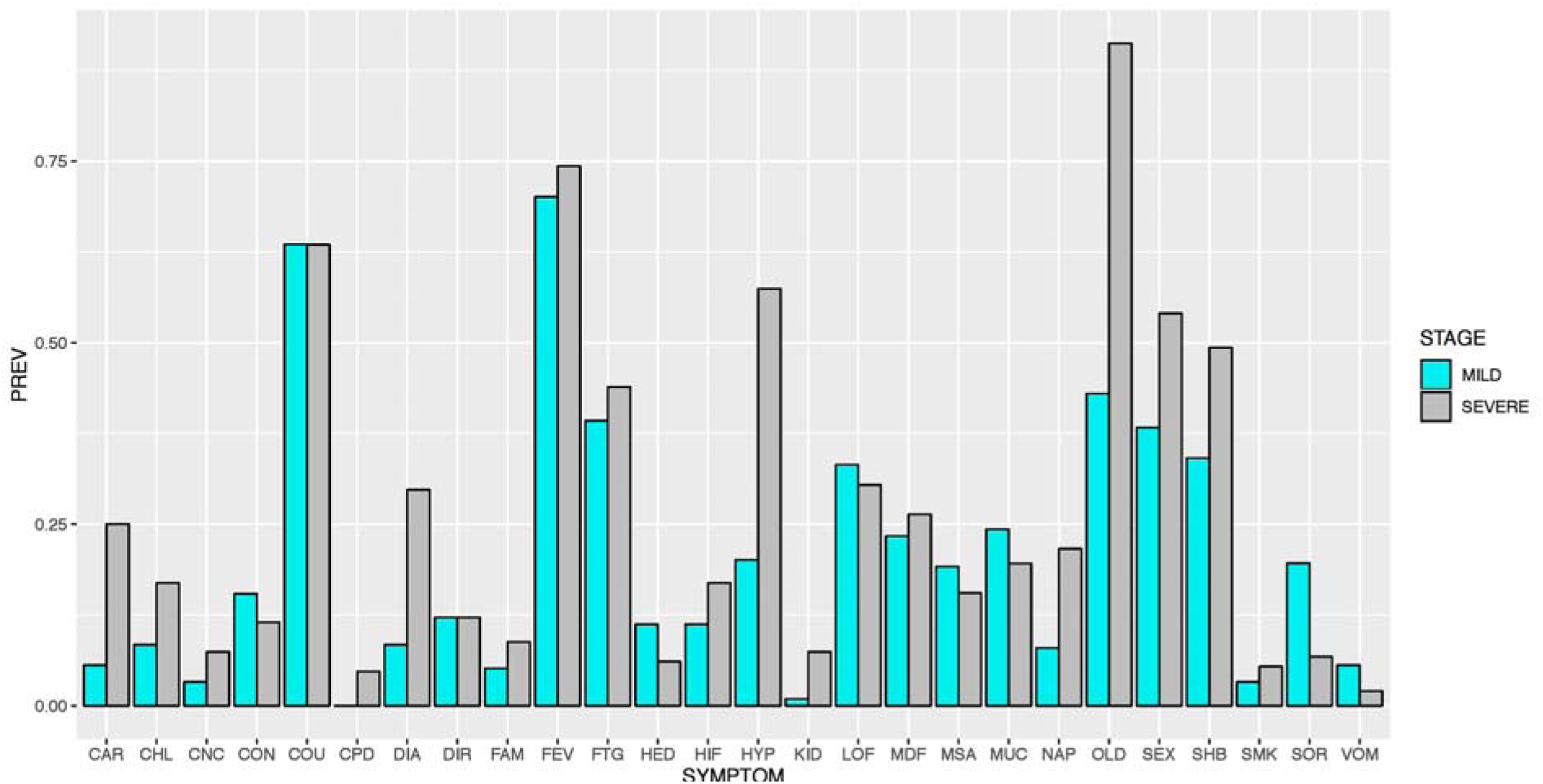
Symptom Features Comparison between Non-severe and Severe Types. Note: symptom features were binary, so Y-axis was the prevalence of positives.

**Fig. 2.**
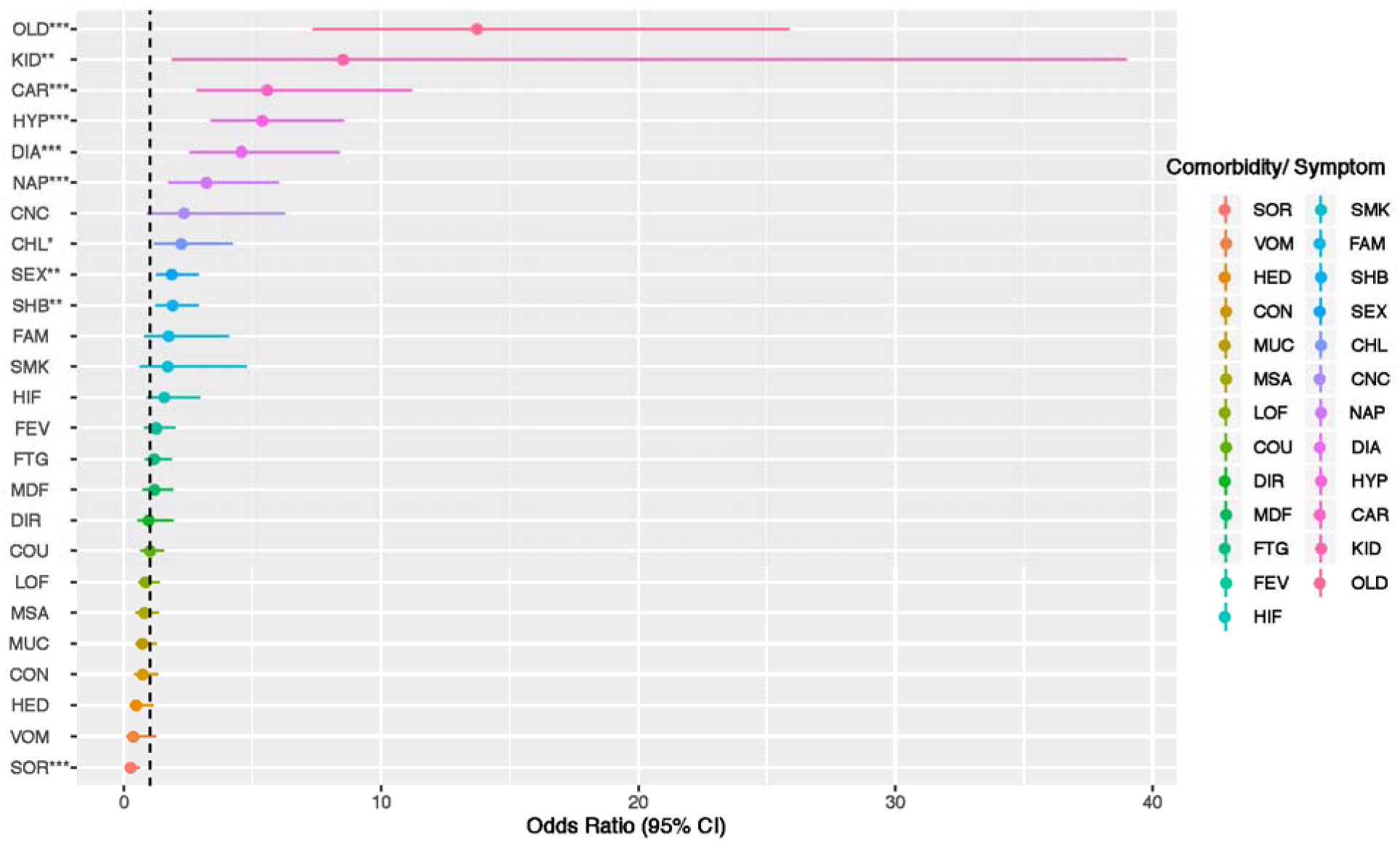
Forest Plot of Symptom Features between Non-severe and Severe Types. Note: *** p<0.001; ** p<0.01; * p<0.05 from the 2×2 contingency table for each feature. COPD was removed from the list because only severe type COVID-19 patients showed comorbidity of COPD. The threshold for a feature to be “positively” or “negatively” associated with severe COVID-19 was 1 (dashed line), not 0. Forest plot is based on parametric statistical analysis and is irrelevant to random forest, a type of machine learning model used later in this study.

For biochemistry modality features, we compared the actual distributions of these continuous features between non-severe and severe types. The results were demonstrated in Fig. 4. Based on the two-sided Kolmogorov-Smirnov test results, severe and non-severe COVID-19 types differed significantly in most biochemistry features, except platelet (PLT), hemoglobin (HGB), CD3, and CD4. Among all biochemistry features, IL-6, hsTNI, and D-dimer had the most significant differences between non-severe and severe COVID-19 types.

**Fig. 3.**
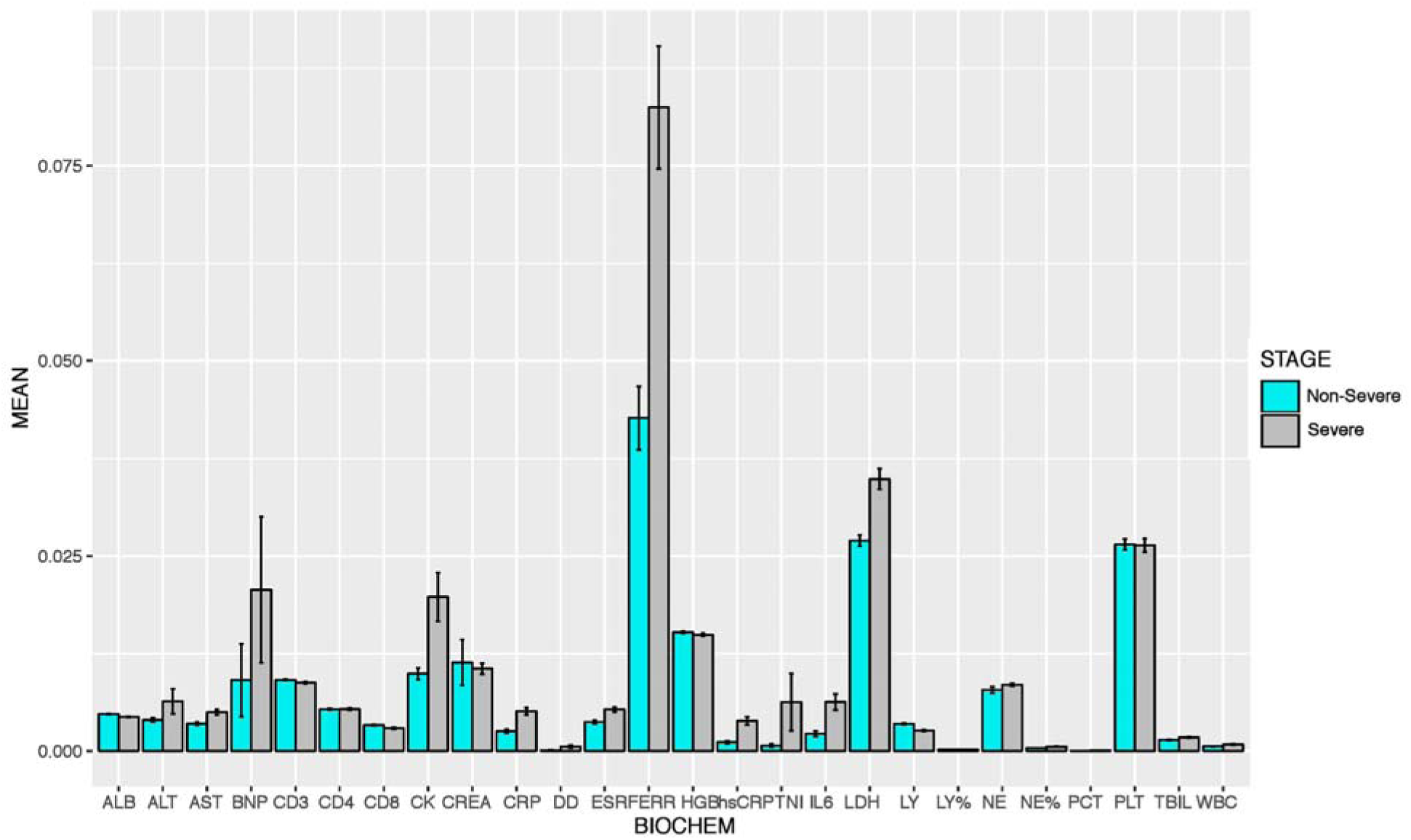
Biochemistry Features Comparison between Non-severe and Severe Types. Note: values shown on y-axis were after feature scaling and were between 0 and 1. Error bars represented standard error (SE) of each biochemistry feature.

In addition, supplementary Fig. 1 showed symptom and biochemistry PCA results between non-severe and severe types. PCA plots reinforced the conclusion that associations among the features were substantially different between the two clinical types. The two types not only had vastly different distributions of features, interrelationships among features were also distinct between the two types.

In conclusion, after extensive clinical feature extraction and data mining, there were strong qualitative and quantitative evidences that non-severe and severe COVID-19 types differed substantially with regard to comorbidities, symptoms, and blood biochemistry. These findings paved the way toward an effective machine learning (ML) classifier to accurately differentiate these two types in clinical practice.

### Clinical Type Classification via Machine Learning (ML): Comorbidity and Symptom Modality

We first explored whether the relatively simple binary symptom features could provide accurate insights on COVID-19 severity. Model performance was summarized in Table 1 upper section. Based on 100 independent runs, the RF model reached an average of >99% and 92% accuracy for training and testing sets, respectively (Table 1). AUC was 90.2% (82.9%-97.6%) based on the receiver operating characteristic (ROC) curve (Fig. 4 left panel). The model performed better in detecting true positives (i.e., severe type) than true negatives (i.e., non-severe type). In other words, symptom features alone in RF models almost never falsely predicted severe case as non-severe case, but with a higher chance to predict non-severe case to severe case. In clinical practice, this would be a lesser concern, as false positive (failed to detect mild type) would be more tolerable than false negative (failed to detect severe type).

**Table 1.**
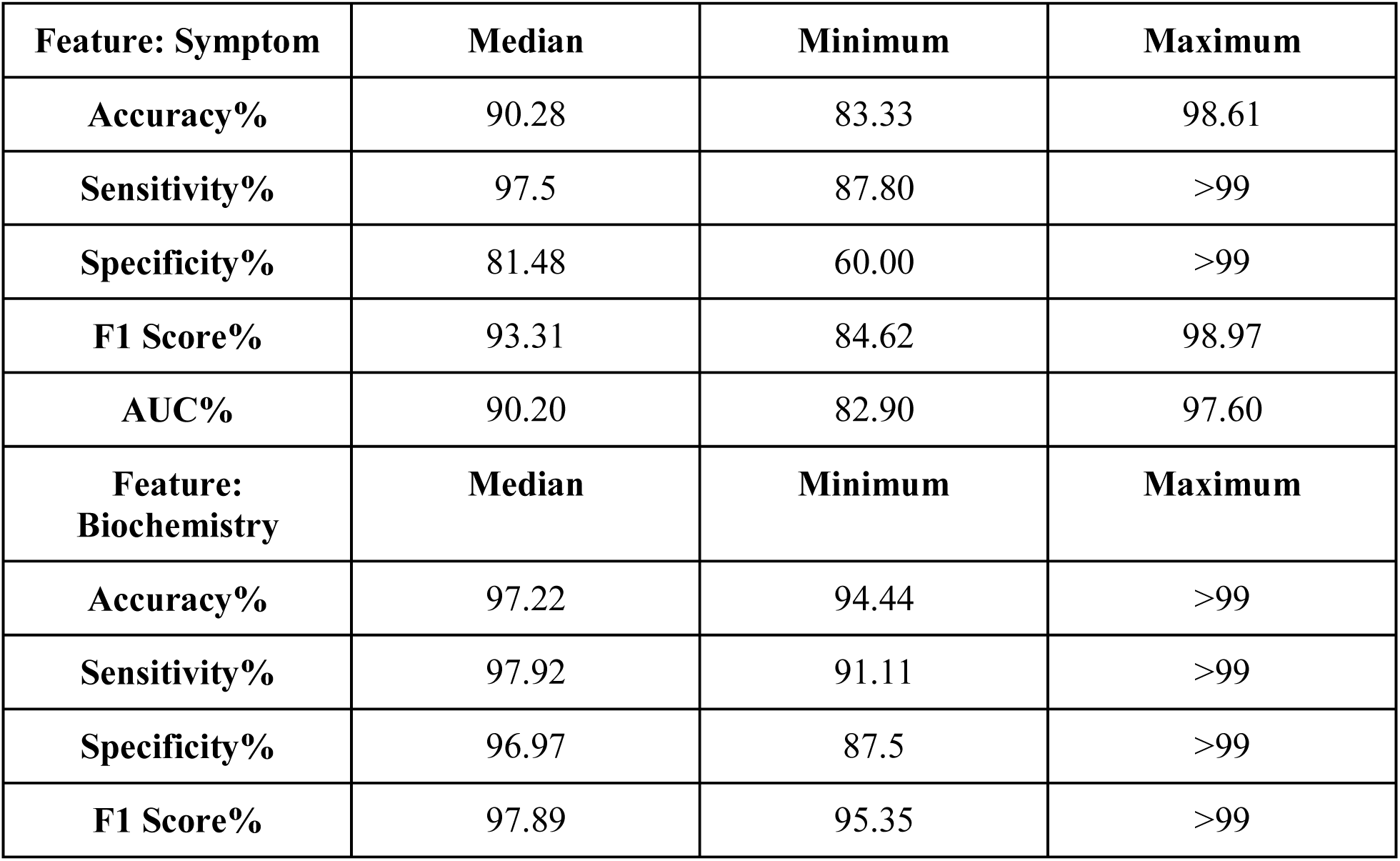
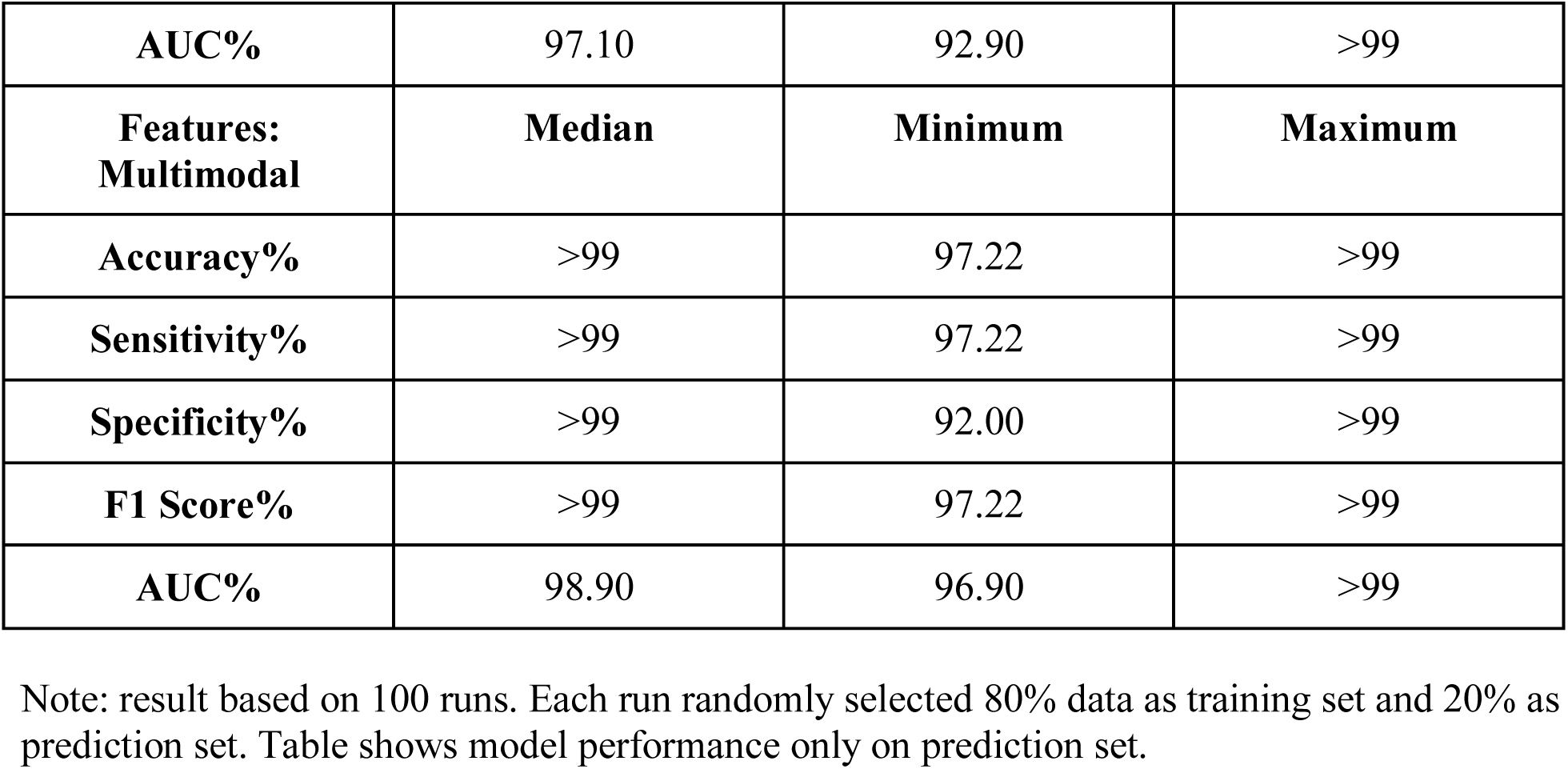
Random Forest Model Prediction Performance with Multimodal Features.

| **Feature: Symptom** | **Median** | **Minimum** | **Maximum** |
| --- | --- | --- | --- |
| **Accuracy%** | 90.28 | 83.33 | 98.61 |
| **Sensitivity%** | 97.5 | 87.80 | >99 |
| **Specificity%** | 81.48 | 60.00 | >99 |
| **F1 Score%** | 93.31 | 84.62 | 98.97 |
| **AUC%** | 90.20 | 82.90 | 97.60 |
| **Feature: Biochemistry** | **Median** | **Minimum** | **Maximum** |
| **Accuracy%** | 97.22 | 94.44 | >99 |
| **Sensitivity%** | 97.92 | 91.11 | >99 |
| **Specificity%** | 96.97 | 87.5 | >99 |
| **F1 Score%** | 97.89 | 95.35 | >99 |
| **AUC%** | 97.10 | 92.90 | >99 |
| **Features:**<br>**Multimodal** | **Median** | **Minimum** | **Maximum** |
| **Accuracy%** | >99 | 97.22 | >99 |
| **Sensitivity%** | >99 | 97.22 | >99 |
| **Specificity%** | >99 | 92.00 | >99 |
| **F1 Score%** | >99 | 97.22 | >99 |
| **AUC%** | 98.90 | 96.90 | >99 |

Note: result based on 100 runs. Each run randomly selected 80% data as training set and 20% as prediction set. Table shows model performance only on prediction set.

**Fig. 4.**
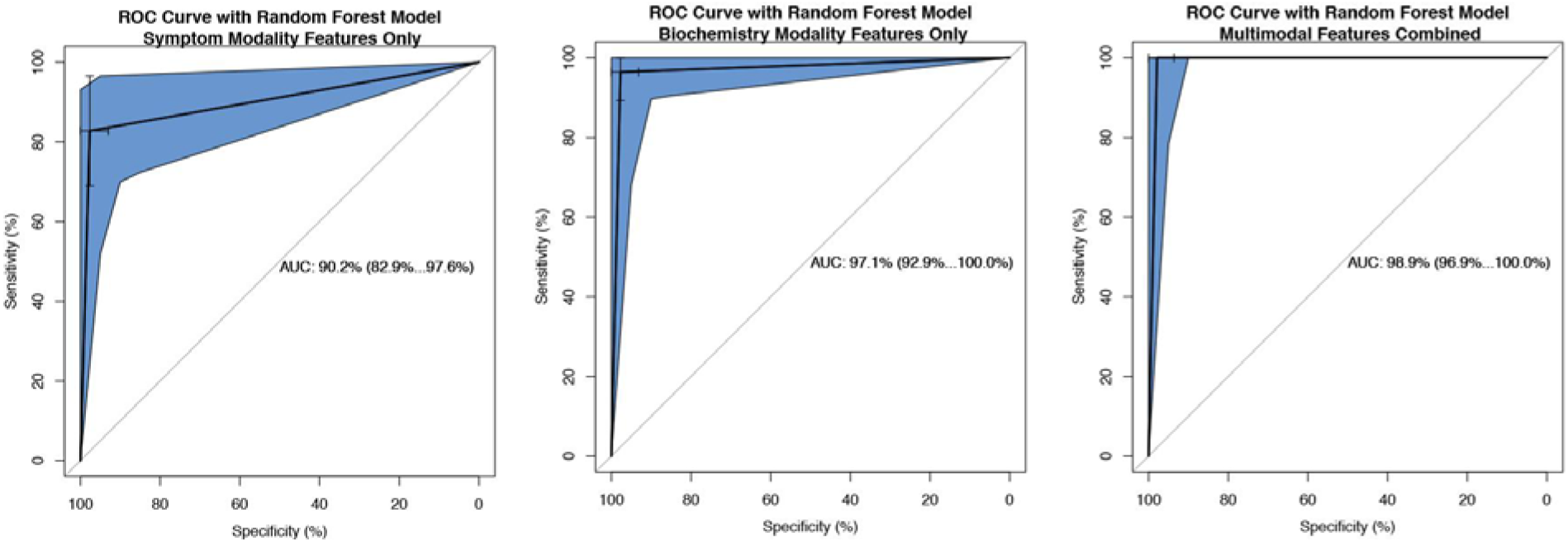
ROC Curve from Random Forest Model Based on Symptom, Biochemistry, and Multimodal Features. Note: panel (A) left showed symptom feature as input alone, panel (B) middle showed biochemistry as input alone, and panel (C) right showed both features combined as input.

Our RF model also provided the major influential features to differentiate COVID-19 types based on contribution to Gini impurity. Top influential features were age, gender, hypertension, diabetes, and cardiovascular diseases, in accordance with existing literature (24). Other important symptom features included fatigue, chest congestion, sore throat, phlegm, and fever. Most of these findings aligned well with our parametric data mining with odds ratio (OR) comparison (Fig. 2, supplementary Table S1) but with much higher accuracy (90% accuracy on prediction set of RF model compared to 68% accuracy from non-ML logistic regression). The only exception was renal disease. While its prevalence was significantly different between the two types, the RF model did not consider it as a major differentiating factor based on Gini impurity (Supplementary Table S1). Clinically, elderly male patients with pre-existing comorbidities, especially hypertension, diabetes, cardiovascular diseases were much more vulnerable to COVID-19 and had a much higher risk to develop to severe type (18,20). Therefore, we suggested using COVID-19 patients’ comorbidity and symptom features as the first round of evaluation of severity with reasonably high accuracy.

### Clinical Type Classification via ML: Biochemistry Modality

Similar to symptom modality, RF model achieved an excellent performance in differentiating non-severe and severe types using 26 features from biochemistry modality. On average, the RF model achieved >99% and >95% accuracy for training and testing sets, respectively. Sensitivity, specificity, and F1 scores were all above 95%, using only 8 trees in the RF model (Table 1, middle section). AUC was 98% based on ROC curve (Fig. 4 middle panel). Though this study focused on ML methods, we evaluated model performance of non-ML logistic regression in supplementary Table S3 as a reference point to show the improvement that state-of-the-art ML models could achieve.

Top differentiating features in biochemistry modality were D-dimer (DD), high sensitivity troponin I (hsTNI), neutrophil (NE), interleukin-6 (IL-6), lactate dehydrogenase (LDH), and high sensitivity c-reative protein (hsCRP). The clinical interpretation of their important role was that severe COVID-19 patients had more intensive immune response and hyperinflammation, such as cytokine storm syndrome with substantially increased IL-6 (25). Research also showed that SARS-CoV-2 was able to infect many organs other than lungs and induce dysfunction of these organs, including heart (26,27). Increasing hsTNI was a sign of heart tissue damage from SARS-CoV-2 infection (28). In addition, severe COVID-19 patients might have formed microthrombosis which induced higher D-dimer (18, 20,29-31). Abnormal level of neutrophils could be responsible for cytokine storm and ARDS in severe COVID-19 patients (13,32). hsCRP, a biomarker of acute inflammation, cardiovascular disease, and ischemic events, was also confirmed as the major contributing factor of COVID-19 mortality (18). LDH was a biomarker of tissue damage and was used to predict the clinical course of COVID-19 patients (42). These findings added further clinical insights in how multiple organs and systems, not just lungs, responded to SARS-CoV-2 infection in different clinical types (33-35).

Therefore, RF models developed in this study provided both high accuracy and valuable insights to identify clinical differences between COVID-19 types as well.

### Clinical Type Classification via ML: Multimodal Features

Based on the success of single modality RF models, we further developed a multimodal RF model that incorporated features and insights from both modalities. Instead of putting every feature in each modality, we only selected top 5 features from symptoms and top 5 features from biochemistry modalities. The results were encouraging and promising: these top 10 features out of a total of 52 features from both modalities achieved >99% in every model performance metric, including accuracy, sensitivity, specificity, and F1 score (Table 1, 3rd section). AUC was >99% as well (Fig. 4 right panel). Therefore, we suggested a two-step evaluation process of COVID-19 patient’s severity in clinical practice. Biochemistry and multimodal features such as a combination of symptom and biochemistry would serve as a more robust second-round confirmation after the first round of initial screening based on binary comorbidity and symptom features.

These findings reinforced our argument that SARS-CoV-2 attacked multiple organs and systems, and the human body reacted in a unity against its invasion. Different clinical features (e.g., comorbidity, symptom, and biochemistry) complemented each other to provide a more comprehensive characterization of human body as a united entity, not just respiratory system, reacted to SARS-CoV-2 invasion (35). In addition, the decent model performance promised the feasibility of multimodal clinical data mining in detecting and differentiating non-severe from severe COVID-19 patients. Our work would help effectively optimize healthcare operation during the pandemic and avoid overloading the healthcare system (7).

## Discussion

This study provides a breakthrough in combining the power of multiple clinical features from different modalities to differentiate COVID-19 clinical types via machine learning techniques. Practically, it enables delivering a more effective and efficient COVID-19 clinical type diagnostic decision support system. It helps develop optimal treatment plans for the individual patient, for example, sending to a mobile cabin hospital or admitting to a hospital with ICU (8). In addition, it will enable triaging and more effectively optimize the healthcare system resources and staffing. Doing so will substantially reduce the risk of overloading the healthcare system by admitting all COVID-19 patients into the hospital, decrease potential healthcare-associated infections, and improve clinical outcome for the patients, especially during this COVID-19 pandemic (7).

In addition to accurately detecting vulnerable COVID-19 patients who are likely to be in severe type, this study also provides clinical insights on why these patients may have been in severe type. Machine learning (ML) models work directly with data and therefore are generally not good at providing clear interpretations. In this study, we combine the power of both hypothesis-driven and data-driven ML models. The most contributing comorbidity, symptom, and biochemical features help predict and explain potential COVID-19 clinical courses and prognosis. Our research echoes recent studies that characterize and predict clinical course, critical illness and mortality of COVID-19 patients (13,18,20). In particular, another decision tree-based algorithm (XGBoost) showed promising performance in predicting mortality of CoVID-19 patients (18). RF was technically similar to XGBoost and our results were consistent to identify the key differentiating biochemistry features: LDH, hsCRP, and lymphocyte.

A continuous-valued risk score calculator for predicting risk of transitioning to critical type (an even more severe type which requires ICU, invasive ventilator, or ECMO, and has a mortality rate as high as 50%) has been developed for COVID-19 patients (20). As a comparison, although our RF model predicts a 0-1 binary outcome for non-severe and severe type patients, the internal RF modeling process through decision tree approach actually calculates an intermediate score between 0 and 1. By using a cut-off threshold, the RF model reports a final dichotomized 0-1 outcome. Therefore, our analytical framework can be readily adjusted to provide a continuous risk score for clinical evaluation and triaging of COVID-19 patients as well, if needed.

Many severe COVID-19 patients present symptoms in lungs, especially ground glass opaque (GGO), which can be detected by biomedical imaging techniques such as CT. However, a major clinical challenge of COVID-19 lies in the asymptomatic patient problem, thus making it far worse than other coronavirus epidemics including SARS and MERS. These patients showed little if not none of classic symptoms related to viral pneumonia, presented no GGO, yet they are almost as capable of transmitting the virus as symptomatic patients (4-6). We suggest that the term “asymptomatic” may be due to lack of a comprehensive evaluation and understanding of this novel pathogen and hosts’ pathophysiology, and not truly “asymptomatic”. By more extensive data mining we show that non-severe COVID-19 patients have many symptoms differently distributed than severe patients. Our study provides an alternative route to detect non-severe COVID-19 patients and complement current biomedical imaging procedures.

The next step of this study is to further include biomedical imaging modality. A technical barrier is that CT scan is a high-dimensional feature set while symptom and biochemistry have relatively low dimensionality. Therefore, CT scan, at its original form of imaging, cannot be effectively combined with other modalities. We will evaluate the feasibility of using convolution neural network (CNN, another time of ML technique) first to reduce feature space in CT scan and extracting a fully connected layer in CNN as a representation of CT scan feature. A fully connected layer is a 1-dimensional vector and has the same dimensionality with the other two modalities. Therefore, in theory we would be able to further combine CT scans with other clinical features and investigate the association between these features with regard to COVID-19.

COVID-19 is a complex disease where the pathogen not only attacks the respiratory system but other organs and systems that have ACE-2 receptors as well (35,36). Our findings reveal the complicated pathological, physiological, and immunological responses to SARS-CoV-2 invasion and shed light in understanding the complex interactions between the virus and human body. Though the multimodal data mining and ML framework is developed with severe/non-severe COVID-19 data, we suggest that the end-to-end framework is applicable to many disease systems where multimodal inputs are common, including demographic information, comorbidity, biochemistry, imaging, and -omics data. Having a more holistic viewpoint and approach will enable us to understand and respond to these emerging diseases, especially the unprecedented COVID-19, more readily in the field. We will further explore this analytical framework, and transfer insights for future clinical studies such as differentiating healthy, non-COVID viral pneumonia, non-severe, and severe COVID-19 patients.

In this study, we recruited participants from a single hospital in Wuhan, the first epicenter of COVID-19. There will inevitably be selection bias, as currently the ethnicity group is limited to Chinese participants. Therefore, we want to inform our colleagues across the world and see whether different demographic backgrounds influence feature distributions between non-severe and severe types in the patients. Our findings have already been independently identified in COVID-19 patients across ethnicity groups (21, 28). Another pitfall we should be aware of is that comorbidities may be the consequence of SARS-CoV-2 invasion, or risk factors that increase the risk of infection. Although the participants’ comorbidity, symptom, and biochemistry were evaluated upon admission to hospital, they could have been exposed to the pathogen long before hospitalization, given the long “asymptomatic” type of COVID-19. For example, it is unclear whether kidney damage in a patient is a risk factor to induce severe type COVID-19, or SARS-CoV-2 attacks the kidney and causes kidney damage (35). The causal relationship needs to be more systematically evaluated with carefully designed prospective cohort studies. Nevertheless, in clinical practice, observing renal diseases in COVID-19 patients would trigger an alarm of clinical course to severe type, and inform clinicians to take actions to prevent acute kidney failure and even death.

Additionally, different subtypes of the virus, their specific pathogenicity and virulence, and host-pathogen interactions, should also be taken into consideration when conducting and comparing studies across different regions of the world. The other factors that this study did not include are behavioral and societal aspects, for instance, whether and how utilizing mobile cabin hospitals to treat non-severe type patients reduce the rate of transition to severe type. COVID-19, like all other infectious diseases, has individual clinical, epidemiological, behavioral as well as societal factors during its epidemic. Therefore, we will also explore cross-scale individual clinical course and population-level epidemics in future studies.

## Materials and Methods

### Data Source and Clinical Feature Extraction

In this study, we recruited 362 COVID-19 patients, including 214 non-severe and 148 severe patients, from Wuhan Union Hospital affiliated to Tongji Medical School, Huazhong University of Science and Technology, China. Definitions of non-severe and severe cases were mainly adopted from the official COVID-19 Diagnosis and Treatment Plan from the National Health Commission of China and consulted guidelines from American Thoracic Society as well (9-11). Patients in severe type should present any one of the following features: 1) respiratory rate > 30 breaths per minute; 2) oxygen saturation < 93% at rest; or 3) arterial oxygen partial pressure (PaO_2_)/fraction of inspired oxygen (FiO_2_) < 300mmHg (40kPa). Each COVID-19 patient was confirmed by two independent qRT-PCR tests before admitted to this study. All patients signed informed consent forms before participation. Symptoms were evaluated and blood samples were drawn upon admission. No pediatric patients younger than 18 years old were admitted in this study. This study was evaluated and approved by the IRB committee of Union Hospital, Wuhan, China (approval number: 2020-IEC-J-345).

Patients’ de-identified clinical information include two major modalities of features. The first modality was a total of 26 pre-existing comorbidities and clinical symptoms, colloquially referred to as symptom features hereinafter. These features included gender, age (dichotomized as elder and young using 50yr as a cut-off point), hypertension, coughing, different types of fever, etc. A detailed description of these 26 features was provided in supplementary table S1. All symptom features were coded as 0-1 binary variables.

In addition, we also collected patients’ blood samples and performed blood chemistry testing. After initial screening, we excluded some features with too many missing data such as calcitonin. Oxygen saturation and PaO2, the severe and non-severe type defining features, according to the Diagnosis and Treatment Plan (9), were also excluded. There were 26 biochemistry features in this study, including IL-6, hemoglobin, and various lymphocytes. A detailed description and units of these biochemistry features were provided in supplementary table 2. All these biochemistry features were continuous features, different from the binary features in the symptom modality.

### Data Mining on Multimodal Clinical Features

First, we conducted data mining on the multimodal COVID-19 data. The original dataset had approximately 5% missing data and we used predictive mean matching (PMM) to impute the original data. PMM was a commonly used computational method to handle missing data. To evaluate the effectiveness of PMM, we used a subset of the original dataset with no data missing, randomly dropped 5% data to simulate potential data loss, re-extrapolated the data with PMM, and evaluated the mean square root error (RMSE) between the original and imputed datasets. The RMSE was less than 0.05, indicating the extrapolation was feasible and reliable. The imputed data were then passed on to successive data mining and machine learning steps.

For the 0-1 binary features in symptom modality, we calculated the prevalence of each feature, i.e., number of positives over the number of patients in each type, non-severe and severe. Z-test was then applied to investigate whether there was a statistically significant difference of prevalence of any binary feature between the two types. In addition, a forest plot of odds ratio (OR) and its 95% confidence interval (CI) of symptom features between severe and non-severe COVID-19 types was developed.

For the continuous features in biochemistry modality, we characterized and contrasted the distribution of each feature in both types. Because most features were not normally distributed, we applied a two-sided Kolmogorovs-Smirnov test instead of Student’s *t*-test for each feature and investigated whether these features distributed differently between the two types. Additionally, principal component analysis (PCA) was applied to help visualize the distinction in feature distributions and associations between the two clinical types.

### COVID-19 Clinical Type Classification via Machine Learning

Traditional hypothesis-driven parametric models, such as logistic regression, relied heavily on human decisions of how features interact with each other (i.e., interaction terms in logistic regression model), which might not reflect the underlying medical reality. In addition, these models had strict prerequisites to perform correctly, for instance, normality of residuals, homoscedasticity, and independence of input features. Initial exploratory analyses showed that input features in both symptom and biochemistry modalities were non-normality and high collinearity among the features. Another technical challenge to logistic regression in this study was a mixture of binary symptom and continuous biochemistry variables from two modalities.

Bearing these problems, logistic regression would not be a preferable modeling approach to accurately classify and predict COVID-19 clinical types. Our exploratory analysis showed that logistic regression could only achieve 68% and 77% predictive accuracy on an 80-20 training-prediction split, using symptom and biochemistry features, respectively (supplementary Table S3). Thus, hypothesis-driven models such as logistic regression were less feasible in clinical settings requiring high accuracy, sensitivity, and specificity to differentiate COVID-19 types.

On the other hand, state-of-the-art machine learning (ML) classification models worked directly with the data to avoid human bias. In addition, ML models did not have restrictions on how input data should be distributed or related. Therefore, ML classification would be a more appropriate modeling approach to predict COVID-19 clinical type with complicated data structure in this study. Therefore, we developed an end-to-end ML framework to accurately predict COVID-19 patient’s clinical type based on symptom and/or biochemistry modality features. We built random forest (RF) classification models, as RF was able to provide excellent interpretability of input variable’s relative importance to support clinical decision-making. RF was a widely used ML model based on decision theory and decision tree approach. The internal validation process through bagging made RF especially accurate and reliable. RF was also robust against data loss and data unbalancing, e.g., more non-severe type patients than severe patients in our study (37-41). There were other types of ML classification methods though, for example, k-nearest neighbor, artificial neural network, and naive Beyes. However, the major goal of this study was not to compare performances of different ML models, we focused on RF to deliver the most accurate classification possible.

We assigned severe cases as "positive" and non-severe as "negative" in the classification. The goal of ML classification through RF was to accurately predict the patient’s COVID-19 type, either "positive" (severe) or "negative" (non-severe), based on features from different clinical modalities. In this part of study, we first use a single modality of features as input. The detailed RF modeling and validation process were provided in supplementary material and method. We trained the model 100 independent times, each time with a randomly selected set of 80% data for training and the remaining 20% for prediction. Hyperparameter number of trees (ntree) in the RF model was set at a very low value (ntree=8) to avoid potential overfitting issues (38). Important ML performance metrics, including accuracy, sensitivity, specificity, F1 score, and area under curve (AUC) value based on receiver operating characteristic (ROC) curve were computed for the prediction set. In addition, RF was able to evaluate input variables’ importance to differentiating the two types based on their contributions to Gini impurity (40). We further quantified input features’ relative importance, identified top contributing features, and explored their clinical relevance and interpretability to COVID-19. The most important features to differentiate COVID-19 clinical types were also cross-checked with our results from exploratory data mining, including prevalence of symptom features and distribution of biochemistry features.

### COVID-19 Clinical Type Classification with Multimodal ML

In addition, we explored whether and how combining features across modalities improved classification performance. In non-ML methods, it would be difficult to combine 0-1 binary inputs with continuous inputs. However, this challenge was non-existent in ML models because ML models worked directly with data without *a priori* assumptions of the data structure.

We developed another RF model that incorporated features from both modalities. The modeling process was exactly the same as using single modality. Nevertheless, instead of putting every feature into the model, we selected top 5 features from each of the two modalities as new inputs. These top features were identified from the Gini impurity of single modality RF models (supplementary Table S1 and S2). We explored whether a few important features from different modalities could perform sufficiently well to address the clinical challenge of differentiating non-severe and severe COVID-19 patients.

All statistical analyses and ML models were built in *R* 3.5.0. and Python 3.7 with additional supporting packages. The codes and fully de-identified data would be freely available on GitHub (https://github.com/forrestbao/corona/tree/master/blood).

## Data Availability

The codes and fully de-identified data would be freely available on GitHub.

https://github.com/forrestbao/corona/tree/master/blood

## Acknowledgments

This work is a tribute to the frontline clinicians and supporting staff who devoted their lives to combating this COVID-19 pandemic.

## Funding

this study was jointly supported by the National Science Foundation for Young Scientists of China (81703201), the Natural Science Foundation for Young Scientists of Jiangsu Province (BK20171076), the Jiangsu Provincial Medical Innovation Team (CXTDA2017029), the Jiangsu Provincial Medical Youth Talent program (QNRC2016548), the Jiangsu Preventive Medicine Association program (Y2018086), the Lifting Program of Jiangsu Provincial Scientific and Technological Association, and the Jiangsu Government Scholarship for Overseas Studies.

## Author contributions

Y.C. designed the study. L.O. and J.L. derived and processed data. L.O., F.S.B., Q.L., L.H., B.Z, J.L., and S.C. interpreted results. M.X. and S.C. performed analyses. Y.G. and S.C. supervised. S.C. developed the manuscript. Y.C., L.O., and F.S.B. contributed equally.

## Competing interests

the authors declare no competing interests in this study.

## Data and materials availability

de-identified clinical data and codes in this study sare openly available on GitHub (https://github.com/forrestbao/corona/tree/master/blood)

## Supplementary Method: Random Forest Machine Learning Model Formulation

In each run, we randomly selected 80% of all data as the training set to train the RF model. These 80% data included both severe and non-severe cases, i.e., both positives and negatives. We also ensured that the distributions of positives and negatives in the training set was similar to those in the complete data. Once the model was developed, the remaining 20% data would be fed into the developed model to evaluate its performance on unseen prediction data. This prediction process was crucial to ensure that the ML model was not over-fitting, i.e., the model worked extremely well on existing training data but poorly on unseen real-world data. We then constructed the 2×2 confusion matrix to evaluate the model performance on prediction data. The 2×2 confusion matrix had four elements, true positive (TP, model correctly identified severe type), true negative (TN, model correctly identified non-severe type), false positive (FP, model incorrectly identified non-severe type as severe type), and false negative (FN, model incorrectly identified severe type as non-severe). Then, important ML model performance metrics were computed, including model accuracy, sensitivity, specificity, and F1 score, etc. Among these performance metrics, accuracy and F1 score evaluated overall performance of the model, sensitivity (also known as true negative rate, TNR) emphasized FN, and specificity (also known as true positive rate, TPR) emphasized FP. Our RF model aimed to increase TP and TN while simultaneously reducing FP and FN. In the other words, an ideal ML model should have both high sensitivity and high specificity. The highest possible value for these metrics was 1 (100%), indicating the model could correctly distinguish all positive (severe) types from negative (non-severe) types. In this study, we run this modeling and predicting process 100 times to evaluate how system stochasticity influenced the RF model and whether the RF model performance was robust. In each of the 100 runs, a different set of randomly selected 80% data were used to train the model and the remaining 20% to predict and evaluate the model performance. We reported maximum, minimum, and median values of performance metrics (accuracy, F1 score, AUC, etc.).

**Supplementary Fig. S1.**
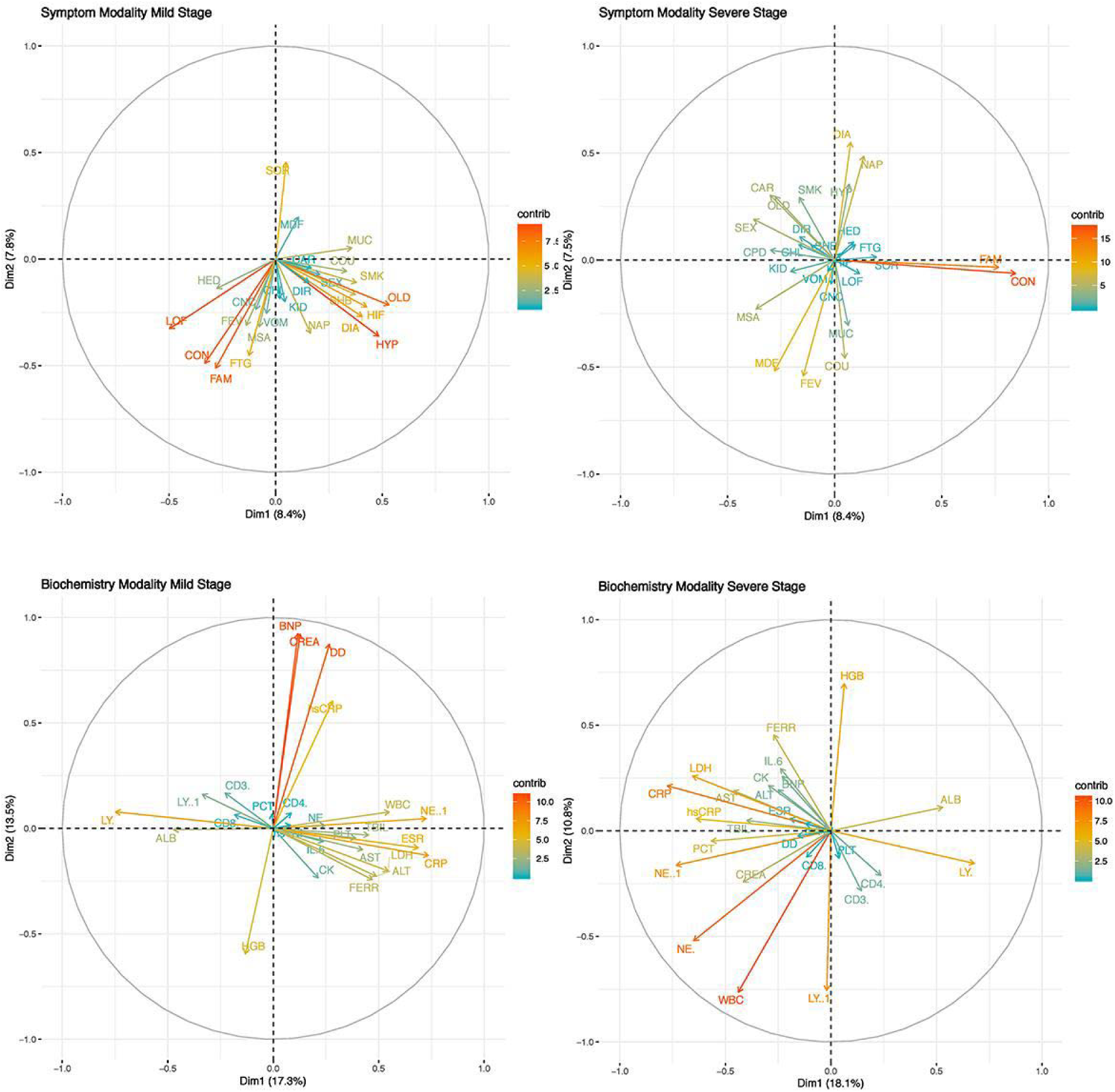
PCA Plot for Features in Symptom and Biochemistry Modalities between Clinical Types. Note: upper section: symptom modality; lower section: biochemistry modality; left section: non-severe (mild) COVID-19 type; right section: severe COVID-19 type.

**Table S1.** Comorbidity and Symptom Features.

| Abbrev. | Health Condition/Symptom | Random Forest Gini Impurity (%) | Logistic Regression Coefficient | Note |
| --- | --- | --- | --- | --- |
| **OLD** | Elderly | 24.94 | 1.85 | Age>50 as elderly (OLD=1) |
| **HYP** | Hypertension | 14.43 | 0.63 | Diastolic>90 or systolic>140 |
| **CAR** | Cardiovascular diseases | 8.57 | 0.75 |  |
| **SEX** | Biological gender | 7.79 | 0.63 | Male=0, female=1 |
| **DIA** | Diabetes | 6.73 | 0.39 | Type 2 diabetes only |
| **FTG** | Fatigue | 6.33 | 0.32 | Subjective, self-reported |
| **SHB** | Chest congestion | 6.29 | 0.28 |  |
| **SOR** | Sore throat | 5.92 | -0.9 |  |
| **MUC** | Phlegm | 5.63 | -0.58 |  |
| **FEV** | Fever (any) | 5.45 | -0.91 | >37C (>98.6F) |
| **COU** | Coughing | 5.41 | -0.05 |  |
| **MSA** | Muscle ache | 5.39 | -0.58 |  |
| **NAP** | Loss of appetite | 5.22 | 0.77 |  |
| **CON** | Contacting COVID-19 patients | 4.33 | -0.28 |  |
| **MDF** | Medium fever | 4.31 | 1.44 | 38.1-39C (100.5-102.2F) |
| **LOF** | Low fever | 4.29 | 1.3 | 37.1-38C (98.7-100.4F) |
| **CHL** | Chilling and shaking | 4.22 | 0.91 |  |
| **DIR** | Diarrhea | 3.86 | -0.45 |  |
| **HIF** | High fever | 3.85 | 1.57 | >39C (>102.2F) |
| **VOM** | Vomiting | 2.79 | -1.7 |  |
| **KID** | Renal diseases | 1.84 | 1.73 |  |
| **HED** | Headache | 1.8 | 0.07 | Any type of headache |
| **CNC** | Cancer | 1.78 | 0.16 | Any type of cancer |
| **FAM** | Family members with COVID-19 | 1.54 | 1.11 |  |
| **SMK** | Smoking history | 1.15 | -0.75 |  |
| **CPD** | Chronic obstructive pulmonary disease (COPD) | 1.08 | 16.28 |  |

**Note:** bold are top ten symptom features critical to differentiate COVID-19 non-severe and severe types from machine learning random forest model. Logistic regression coefficient signs (positive or negative) reveal if the feature increases or decreases risk of developing severe type COVID-19.

**Table S2.** Blood Biochemistry Features.

| Abbrev. | Blood Biochemistry | Importance<br>by Gini<br>Impurity (%) | Logistic<br>Regression<br>Coefficient | Unit and Note |
| --- | --- | --- | --- | --- |
| **DD** | D-dimer | 25.41 | 0.5 | mg/L |
| **hsTNI** | High sensitivity<br>Troponin I | 16.06 | 0.0031 | ng/mL |
| **LDH** | Lactate dehydrogenase | 10.19 | 0.0012 | U/L |
| **NE** | Neutrophil | 10.02 | 0.0044 | 109/L |
| **IL6** | Interleukin 6 | 9.41 | 0.026 | ng/mL |
| **hsCRP** | High sensitivity C-reactive protein | 9.11 | 0.019 | ug/L |
| **ESR** | Erythrocyte<br>sedimentation rate | 7.96 | 0.029 | mm/h |
| **TBIL** | Total bilirubin | 7.43 | 0.018 | umol/L |
| **CD8** | Cluster of differentiation<br>8 | 7.09 | -0.097 | /uL |
| **CK** | Creatine kinase | 6.7 | <0.001 | U/L |
| CRP | C-reactive protein | 6.69 | -0.02 | ug/L |
| FER | Ferritin | 5.86 | <0.001 | ug/L |
| ALT | Alanine transaminase | 5.52 | -0.0037 | U/L |
| CREA | Creatinine | 4.45 | 0.01 | umol/L |
| LY% | Percent of Lymphocyte | 4.4 | 0.91 | % |
| CD3 | Cluster of differentiation<br>3 | 4.06 | 0.051 | /uL |
| ALB | Albumin | 4.02 | 0.015 | g/L |
| NE% | Percent of Neutrophil | 3.94 | 0.51 | % |
| PLT | Platelet | 3.91 | <0.001 | 109/L |
| AST | Aspartate<br>aminotransferase | 3.85 | 0.017 | U/L |
| PCT | Procalcitonin | 3.57 | -0.28 | ng/mL |
| CD4 | Cluster of differentiation<br>4 | 3.51 | -0.056 | /uL |
| LY | Lymphocyte | 3.26 | -0.055 | 109/L |
| WBC | White blood cell | 3.13 | -0.4 | 109/L |
| BNP | Brain natriuretic peptide | 3.07 | 0.0033 | pg/mL |
| HGB | Hemoglobin | 2.98 | -0.024 | g/L |

**Note:** bold are top ten biochemistry features critical to differentiate COVID-19 non-severe and severe types from machine learning random forest (RF) model.

**Table S3.** Logistic Regression Prediction Performance.

| Feature | Symptom | Biochemistry |
| --- | --- | --- |
| Accuracy% | 69.44 | 78.62 |
| Sensitivity% | 65.22 | 78.65 |
| Specificity% | 76.92 | 78.57 |
| F1 Score% | 71.07 | 78.61 |

**Note:** multimodal logistic regression is not performed because two feature modalities are not compatible to combine, i.e., 0-1 binary symptom and continuous biochemistry input data.

## Notes

### Competing Interest Statement

The authors have declared no competing interest.

